# Efficacy of B_12_ fortified nutrient bar and yogurt in improving plasma B_12_ concentrations – results from two double blind randomised placebo controlled trials

**DOI:** 10.1101/2021.01.01.21249112

**Authors:** Chittaranjan Yajnik, Sonal Kasture, Vaishali Kantikar, Himangi Lubree, Dattatray Bhat, Deepa Raut, Nilam Memane, Aboli Bhalerao, Rasika Ladkat, Pallavi Yajnik, Sudhir Tomar, Tejas Limaye, Sanat Phatak

## Abstract

**Background:** Dietary vitamin B_12_ (B_12_) deficiency is common in Indians. It may affect hematologic and neurocognitive systems and maternal deficiency predisposes offspring to neural tube defects and non-communicable disease. Long-term tablet supplementation is not sustainable.

**Objective:** To study efficacy of B_12_ fortified nutrient bar and yogurt in improving plasma B_12_ concentrations in children and adults.

**Methods:** Two double-blind, placebo-controlled randomised directly observed therapy (DOT) trials were conducted: 1. Healthy children (10–13Y) were fed nutrient bar fortified with B_12_ (2 mcg), multiple micronutrients (B_12_ 1.8 mcg) or placebo for 120 days. 2. Healthy adults (18–50Y) were fed yogurt fortified with B_12_ (2 mcg) or Propionibacterium (1×10^8^cfu/g) or placebo for 120 days. B_12,_ folate, homocysteine and hemoglobin concentrations and anthropometry were measured before and post intervention.

**Results:** We randomised 164 children and 118 adults; adherence was 96% and 82% respectively. In children, B_12_ fortified bars increased B_12_ concentrations significantly above baseline (B_12_ alone: median +91 pmol/l, B_12_+ multiple micronutrients: +82 pmol/l) compared to placebo. In adults, B_12_ fortified yogurt increased B_12_ significantly (median +38 pmol/l) compared to placebo, but Propionibacterium did not. In both trials, homocysteine fell significantly with B_12_ supplementation. There was no significant difference in different groups in anthropometry and hemoglobin.

**Conclusions:** B_12_ fortified foods are effective in improving B_12_ status in Indian children and adults. They could be used to improve vitamin B_12_ status in the national programs for children, adolescents and women of reproductive age. They could also be used as over the counter products.

**Brief Highlights:** Vitamin B12 deficiency is common in India. Culturally acceptable fortified foods will help reduce it. We performed two RCTs (children and adults) with B12 fortified nutrient bar and yogurt at near RDA doses and found significant improvement in B12 status. This has important implications for nutritional policy.

## Introduction

Low Vitamin B_12_ (B_12_) status is widely prevalent in vegetarian populations^1^. Though asymptomatic in many individuals, deficiency is associated with haematologic, neurocognitive and cardiovascular manifestations in some^2^. Low maternal B_12_ status during pregnancy is linked to increased risk of neural tube defects (NTDs), preeclampsia, gestational diabetes, preterm delivery, fetal growth restriction, as well as to increased risk of future adiposity and insulin insensitivity in the child^3^.

Only bacteria synthesize B_12_ naturally^4^. Animals get their B_12_ from food and water colonised with B_12_ producing microorganisms. Foods rich in B_12_ include meat, liver, fish, eggs, and milk. Plant foods do not contain B_12_. Low B_12_ status in developing countries is largely attributed to low intake of animal-origin foods, either because of cultural and religious ethical practices (Hindu, Jain and Buddhist), or due to poverty, which precludes expensive non-vegetarian foods from diet^5^. Low B_12_ status is common in Indians, both in those living in India as well as those migrated abroad^6^. This is largely attributable to vegetarianism but not to pernicious anemia (malabsorption due to lack of intrinsic factor). Our research showed that Vit B_12_ deficiency is common in and around Pune, India^6^, despite normal B_12_ absorption in > 85%of the population^7^. B_12_ status improved in community based trials by supplementation with capsules containing 2 mcg B_12_ per day ^8^ and drinking 400 ml of milk daily^9^.

Increasing intake of animal origin foods to improve vit B_12_ status has obvious practical limitations in Indians. Though low dose supplements are successful in a trial setting, they are unlikely to have a big impact in public health owing to poor long-term adherence to medications in asymptomatic individuals. Food fortification may be an effective way to improve B_12_ status in Indians and other vegetarian populations. Fortification of breakfast cereals and milk with B_12_ is common in many countries but not in India. In recent years, probiotics including Propionibacterium have been claimed to improve B_12_ status^10^. This could also be of help in Indians and warrants further research.

As part of our research to improve B_12_ status of the population, we tested two B_12_ fortified food items - i) A nutrient bar in school children, and ii) Yogurt in adults, for their efficacy to improve B_12_ status. We also tested a Propionibacterium species claimed to produce substantial quantities of B_12_ as a separate fortificant for yogurt.

## Methods

### Study Design and Intervention

We conducted two double blind, placebo-controlled, randomized, directly observed therapy (DOT) trials. We excluded those with chronic medical illness, those on vitamin supplementation and those with very low B_12_ concentration (<100 pmol/l) or anemia (hemoglobin <10 g/dl) for ethical reasons and were advised appropriate treatment. Randomization was computer generated, and stratification was based on baseline B_12_ and hemoglobin concentrations (below and above median) to ensure comparable distribution. All participants were instructed not to take any vitamin supplements during the trial. Adherence was calculated for each participant by calculating the number of days of attendance during the trial.

### Nutrient bar trial

This was conducted in a village school (Pabal, Pune District, approximately 70 km from Pune) in children aged 10–13 years. School authorities approved participation in the trial. We arranged a meeting with the school children and their parents to explain the trial and invited them to sign an assent and consent. Included children were randomized to 3 groups to receive one bar per day, fortified with a) 2mcg B_12_ or b) multiple micronutrients (MMN, including 1.82 mcg B_12_ (UNICEF guidelines)^11^, or c) no added micronutrients (placebo). All bars were similar in appearance and taste. The children ate the bars in the school in the morning recess, directly observed by research staff. The duration of supplementation was 120 consecutive school days, from September 2011 to March 2012.

### Yogurt trial

The yogurt study was conducted in volunteers aged 18–50 years at the KEM Hospital, Pune. Hospital staff members and their friends were invited to enroll through notice board advertisement. Eligible participants were randomised into 3 groups to receive 100 gm yogurt fortified with a) 2µg B_12_, b) 1×10^8^cfu/g *Propionibacterium*, and c) without any additions (placebo). The strain used for fortification (Propionibacterium freudenreichi subsp. freudenchii ATCC 6207, GRAS certified) was provided by the National Dairy Research Institute (NDRI, Karnal, Haryana, India) and was recommended because it was known to produce B_12_ in vitro^12^. The three yogurt preparations were similar in appearance, taste, flavor and smell. The yogurt was eaten under supervision for 120 consecutive working days between December 2013 and May 2014.

In nutrient bar trial, informed written assent was obtained from the children, and consent was signed by the parents. In yogurt trial, participants provided written consent. Both trials were approved by KEM Hospital Research Centre Ethics Committee. For the nutrient bar trial, permissions were obtained from the school authorities and the District Health Officer. The trials were registered with the Clinical Trials Registry of India (CTRI/2012/07/002799 - nutrient bar, CTRI/2015/04/005703 -yogurt). Adverse events were recorded.

### Measurements

Height was measured to the nearest 0.1 cm using a stadiometer (CMS Instruments, London, UK) and body weight to the nearest 0.01 kg using an electronic weighing scale (Model no. HD-358, Tanita Corporation, Japan). At baseline, non-fasting venous blood sample was collected in EDTA vacutainers for measurements of hemogram, B_12_, folate and homocysteine. Hemogram was measured on a Beckman Coulter Analyzer (AC.T diffTM, Miami, Florida, USA) on the same day. Plasma aliquots were stored (−70° C) until further analysis. B_12_ was measured by a microbiological assay using a colistin sulfate-resistant strain of *Lactobacillus leichmannii* (CV <8%) ^13, 14^. Plasma total homocysteine (homocysteine) was measured by fluorescence derivative of monobromobiame using HPLC (inter and intra batch CV <4%). Plasma folate was measured by a microbiological assay using a chloramphenicol-resistant strain of *Lactobacillus Casei* (inter and intra CV <8%) ^15,16^. The blood and clinical measurements were repeated at the end of the trial.

### Statistical analysis

Data are presented as median (25^th^ – 75^th^ centile) for continuous variables and as percentages for categorical variables. Skewed variables were log normalized before analysis. Significance of the differences between baseline and end of trial levels of clinical and biochemical measurements were tested by paired t test for continuous variables and by chi-square test for categorical variables. Significance of difference from placebo group was tested by t-test. Analysis was by intention to treat (ITT) method. The participants who were lost to follow-up were analyzed by ‘last observation carried forward’ (LOCF) method.

For children, we calculated sample size required in each group to demonstrate a 50% rise in plasma B_12_ concentrations at 5% significance level and found that 55 individuals allowing a dropout rate of 10% will provide a power of more than 90%. For adults, we calculated sample size required in each group to demonstrate a 30% rise in plasma B_12_ concentrations at 5% significance level and found that 45 individuals allowing a dropout rate of 10% will provide a power of more than 80%.

## Results

### Nutrient bar trial

We approached 180 school children and their parents, of whom 178 (99%) agreed to participate. Fourteen children were excluded: 13 with B_12_ <100 pmol/l and 1 anemic; they were prescribed appropriate treatment (figure -1a). The 164 randomized children (57% girls) were 11.3 (10.9 – 12.0) years old, 139.4 (134.5 – 144.5) cm tall and with a weight of 28.8 (25.2 – 33.4) kg; there was no significant difference between groups. Plasma vitamin B_12_, folate and homocysteine concentrations were comparable. Table-1

**Table 1:**
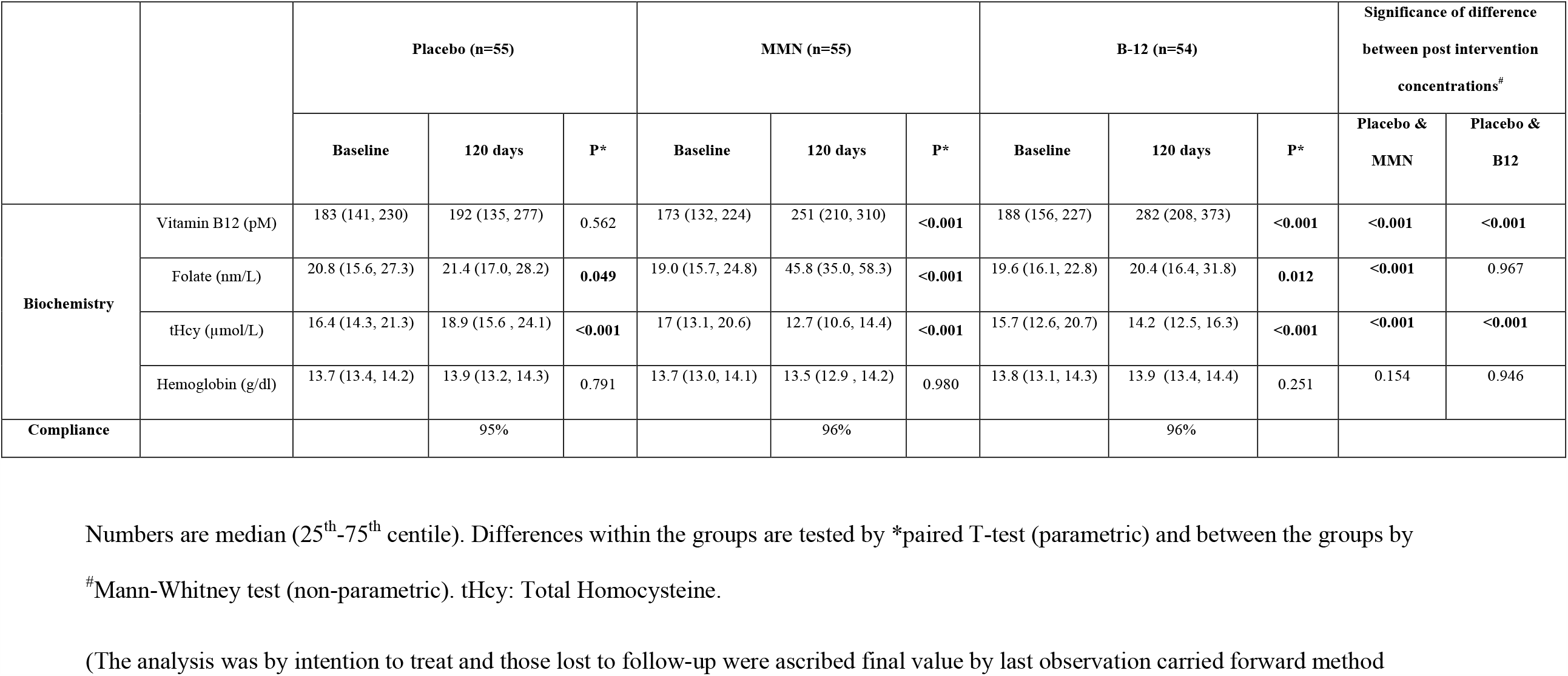
Nutrient bar trial - comparison within and between groups

|  |  | Placebo (n=55) |  |  | MMN (n=55) |  |  | B-12 (n=54) |  |  | Significance of difference between post intervention concentrations<sup>#</sup> |  |
| --- | --- | --- | --- | --- | --- | --- | --- | --- | --- | --- | --- | --- |
|  |  | Baseline | 120 days | P* | Baseline | 120 days | P* | Baseline | 120 days | P* | Placebo & MMN | Placebo & B12 |
| **Biochemistry** | Vitamin B12 (pM) | 183 (141, 230) | 192 (135, 277) | 0.562 | 173 (132, 224) | 251 (210, 310) | <0.001 | 188 (156, 227) | 282 (208, 373) | <0.001 | <0.001 | <0.001 |
|  | Folate (nm/L) | 20.8 (15.6, 27.3) | 21.4 (17.0, 28.2) | 0.049 | 19.0 (15.7, 24.8) | 45.8 (35.0, 58.3) | <0.001 | 19.6 (16.1, 22.8) | 20.4 (16.4, 31.8) | 0.012 | <0.001 | 0.967 |
|  | tHcy (μmol/L) | 16.4 (14.3, 21.3) | 18.9 (15.6, 24.1) | <0.001 | 17 (13.1, 20.6) | 12.7 (10.6, 14.4) | <0.001 | 15.7 (12.6, 20.7) | 14.2 (12.5, 16.3) | <0.001 | <0.001 | <0.001 |
|  | Hemoglobin (g/dl) | 13.7 (13.4, 14.2) | 13.9 (13.2, 14.3) | 0.791 | 13.7 (13.0, 14.1) | 13.5 (12.9, 14.2) | 0.980 | 13.8 (13.1, 14.3) | 13.9 (13.4, 14.4) | 0.251 | 0.154 | 0.946 |
| **Compliance** |  |  | 95% |  |  | 96% |  |  | 96% |  |  |  |

Numbers are median (25<sup>th</sup>-75<sup>th</sup> centile). Differences within the groups are tested by \*paired T-test (parametric) and between the groups by
<sup>#</sup>Mann-Whitney test (non-parametric). tHcy: Total Homocysteine.
(The analysis was by intention to treat and those lost to follow-up were ascribed final value by last observation carried forward method

**Figure 1.**
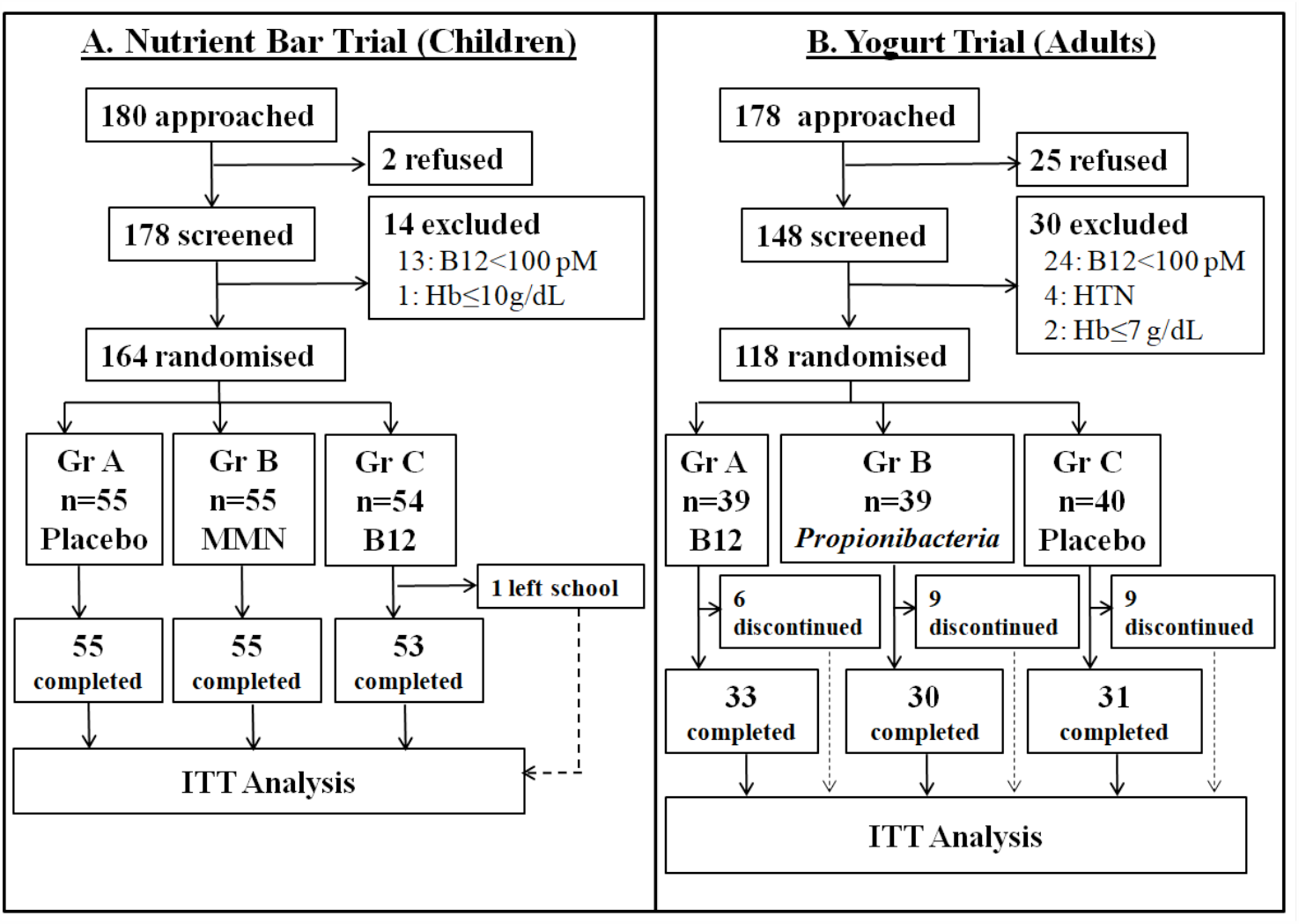
shows flow of participants in two trials. A. Nutrient bar trial was conducted in village school children. B. Yogurt trial was conducted in healthy adult volunteers in an urban setup

After intervention, B_12_ rose significantly by median 91 pmol/l and 82 pmol/l above the baseline in B_12_ alone and B_12_ + MMN group respectively (p<0.001) while there was a no change in the placebo group (figure -2). There was a small rise in folate concentrations in the placebo and B_12_ groups, and substantial rise in the MMN group. Homocysteine reduced by median 1.4 and 3.8 µmol/l in the B_12_ alone and B_12_+ MMN groups respectively (p<0.001), and increased by median 1.9 µmol/l (p<0.001) in the placebo group (figure -2). Hemoglobin concentrations did not change with the intervention. There was an average gain of 4.3 (3.6 – 5.2) cm in height and 2.9 (2.0 – 4.4) kg in weight post intervention but no significant difference between the groups.

**Figure 2.**
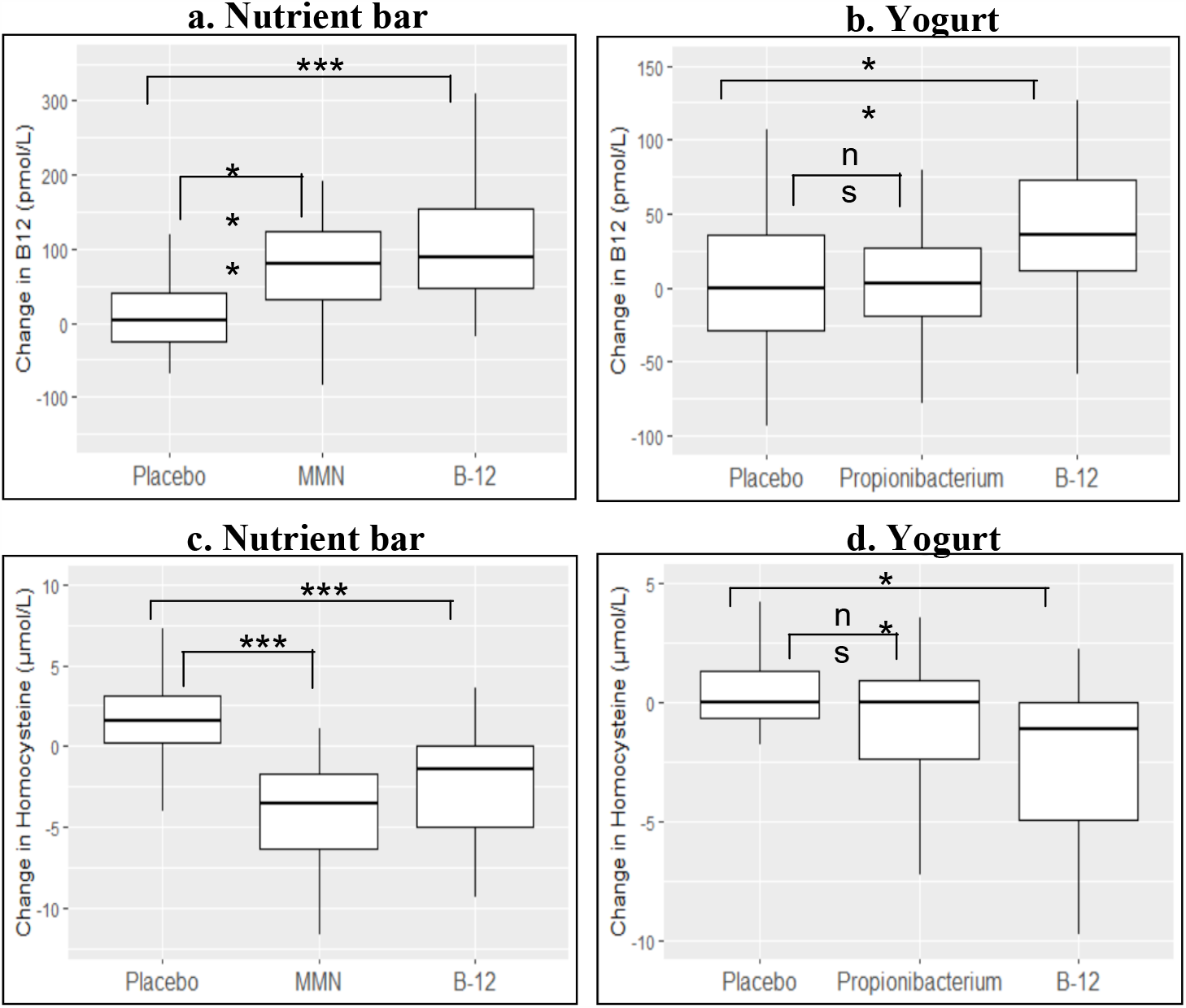
(box plot) shows change in plasma vitamin B12 and homocysteine concentrations after supplementation with fortified food products for 120 days. Differences between the groups are tested by Mann-Whitney test. P-values are given as * <0.05, **<0.01, ***<0.001, ns: non significant. MMN: Multi Micro Nutrient (1.8 mcg vitamin B12), B-12: Vitamin B12 (2.0 mcg), Propionibacterium: *Propionibacterium freudenreichi* (1×10^8^cfu/g)

### Comparison with placebo

The post intervention concentrations of B_12_ and rise above baseline were higher in both B_12_ supplementation groups compared to those in the placebo group. Rise and post intervention concentrations of folate were higher in the MMN group compared to the placebo and B_12_ group. Fall in homocysteine concentrations was significantly greater in both the B_12_ and MMN groups compared to the placebo group (figure -2).

Mean adherence for daily nutrient bar consumption was high and similar in three groups (≥95%). One child dropped out during the trial because of change of school (figure -1a). There were a total of four serious adverse events (hospital admissions for incidental illnesses), none related to the intervention product.

### Yogurt trial

Of 178 adult volunteers, 148 consented to participate. Thirty were excluded: 24 with B_12_ <100 pmol/l, 2 with anaemia and 4 due to medical disorders (figure -1b). The 118 randomized adults (81% women) were 27.0 (27.8 – 35.0) years old, 157.5 (152.8 – 162.5) cm tall, with a weight of 56.5 (49.4 – 64.6) kg and BMI of 22.9 (19.7 – 26.0) kg/m^2^, there was no significant difference between groups. Plasma vitamin B_12_, folate and homocysteine concentrations were comparable. Table-2

**Table 2:** Yoghurt trial: comparison within and between groups

|  |  | Placebo (n=40) |  |  | Propionibacterium (n=39) |  |  | B12 (n=39) |  |  | Significance of difference between post intervention concentrations<sup>#</sup> |  |
| --- | --- | --- | --- | --- | --- | --- | --- | --- | --- | --- | --- | --- |
|  |  | Baseline | 120 days | P* | Baseline | 120 days | P* | Baseline | 120 days | P* | Placebo & Propionibacterium | Placebo & B12 |
| Biochemistry | B12 (pM) | 151 (123, 239) | 167 (120, 234) | 0.670 | 167 (125, 214) | 168 (142, 216) | 0.460 | 142 (118, 185) | 207 (154, 278) | <0.001 | 0.212 | 0.057 |
|  | Folate (nm/L) | 14 (11.0, 22.3) | 18.8 (14.5, 24.4) | 0.062 | 18.1 (13.7, 23.6) | 20.5 (17.3, 28.4) | 0.009 | 16 (12.1, 22.6) | 19 (17.2, 27.8) | 0.001 | 0.104 | 0.994 |
|  | tHcy (μmol/L) | 19.0 (11.9, 34.9) | 19.3 (12.0, 32.6) | 0.675 | 16.9 (12.0, 25.0) | 17.3 (12.1, 23.4) | 0.815 | 22.7 (14.3, 43.5) | 17.3 (12.9, 29.8) | 0.001 | 0.426 | 0.720 |
|  | Hemoglobin (g/dl) | 11.5 (10.0, 12.4) | 11.9 (10.5, 13.0) | 0.015 | 11.2 (9.5, 12.0) | 11.3 (10.1, 12.3) | 0.018 | 11.4 (10.4, 13.4) | 11.6 (10.2, 14.0) | 0.539 | 0.511 | 0.935 |
| Compliance |  |  | 77% |  |  | 84% |  |  | 85% |  |  |  |

Numbers are median (25<sup>th</sup> – 75<sup>th</sup> centile). Differences within the groups are tested by \*paired T-test (parametric) and between the groups by
<sup>#</sup>Mann-Whitney test (non-parametric). tHcy: Total Homocysteine.
(The analysis was by intention to treat and those lost to follow-up were ascribed final value by last observation carried forward method)

After intervention, B_12_ rose significantly by median 38 pmol/l above the baseline in the B_12_ group (p<0.001) and homocysteine decreased by median 2.7 µmol/l (p<0.001) (figure -2). There were no significant changes in B_12_ or homocysteine in the Propionibacterium and the placebo groups. There was a small rise in folate concentrations in all groups, but no significant changes in hemoglobin concentrations and weight in any group.

### Comparison with placebo

The rise in B_12_ concentration and fall in homocysteine concentration from baseline was higher in B_12_ fortified yogurt group compared to the placebo and Propionibacterium fortified groups. The fall in the homocysteine concentration was relatively smaller and not significantly different between groups (figure -2).

During the intervention 6 participants discontinued from the B_12_, 9 from *Propionibacterium* and 9 from plain yogurt group; all for reasons not related to intervention. Ninety four (82%) participants completed the trial (figure -1b). Mean adherence for daily yogurt consumption was moderate and similar in three groups (≥82%). There was one serious adverse event (hospital admission for incidental illness) not related to the intervention product.

## Discussion

Our results show that regular consumption of B_12_ fortified nutrient bar or yogurt for 4 months significantly improved B_12_ status. Our population has high prevalence of B_12_ deficiency, and we used near RDA doses to make it public health relevant. DOT design ensured high adherence and allowed us to explore the full potential of fortification with near RDA doses. The response was proportional to the dose of B_12_ per kg body weight, and therefore, higher in children compared to adults. Placebo comparison helped highlight the specific effects of vitamin B_12_ interventions. Nutrient bar and yogurt are two commonly eaten food items in India, and were well accepted by the participants. Our results make it an attractive public health proposition. The Propionibacterium probiotic at the recommended dose did not influence B_12_ status.

Improvement in B_12_ status reflected in reduction of circulating homocysteine concentrations, indicating an improvement in the methylation status of the body in both the children and the adults. Addition of other micronutrients (folic acid, B6 and B2) to B_12_ had an additional effect on lowering of homocysteine in children. There was no effect on hemoglobin concentration in these trials, which might reflect a requirement for a relatively higher dose of these vitamins for hematological effects. In another study of more severely B_12_ deficient adolescent girls we were able to show improvement in hemoglobin concentration (and peripheral nerve function) with similar daily dose of vit B_12_ but over a longer period of 11 months.^17^ Our results support fortification of commonly eaten food items as vehicles for improving B_12_ status in populations with low B_12_ status who do not have the problem of defective absorption.

There are very few studies of exclusive B_12_ fortification to improve the vitamin status. They have been done in pre-school children (MMN) ^18,19^ and in elderly (B_12_)^20^, the two groups commonly thought to be at risk of low B_12_ status. The vehicles for B_12_ were rice, milk, wheat flour (consumed as bread), common salt and breakfast cereals.^18, 19, 21-23^ In a population with high burden of low B_12_ status, a multipronged food fortification program may be more successful than isolated food product fortification to improve the B_12_ status. In our own and others’ experience, supplementation with tablets in asymptomatic individuals suffers rapid reduction in adherence. Such supplementation also has complex logistic requirements in field operations in large populations.

We performed our investigations in a population with high prevalence of low B_12_ status, making it relevant to many such populations in India and other developing countries.^24^ Participation rates were high. The placebo-controlled, randomized design ensured a causal association for both the trials. Use of DOT approach ensured near complete compliance (≥95%) in children, thus allowing full potential of the intervention to be exploited. Our findings will be directly applicable to vulnerable populations (children, adolescents, young adults and pregnant women) where low B_12_ status may have major implications for growth, development and reproductive health. Possible weaknesses include small sample size and a relatively short period of intervention. Relatively lower adherence in the adults (average 82%) is a reflection of the real-life situation in working middle class and not a reflection on the investigational product. Despite these, there was a rewarding improvement in B_12_ status.

In summary, we used fortified versions of two commonly consumed food items to improve B_12_ status in deficient populations. This approach appears superior to use of vitamin tablets, which has proved difficult to sustain over longer periods of time in asymptomatic population. Both food items additionally provide calories, proteins and other nutrients and therefore could be easily adopted in national programs of feeding malnourished children and adults, as well as in the Mid-day Meal programs in schools. There is a scope to perform a longer trial to investigate benefits to physical, cognitive and reproductive outcomes. Both items could be made at low cost by small-scale home industry promoting local economy. Future trials in community will help estimate effectiveness of these food fortifications in deficient populations.

## Data Availability

Data is available on request from the corresponding author, Prof C S Yajnik. Data requests can be emailed to.

## Acknowledgements

The study was funded by Department of Bioechnology (DBT), New Delhi, India. DSM India kindly provided nutrient bars for the study. Hi Tech Biosciences India Limited, Pune produced yogurt for the trial. We thank the team at Diabetes Unit, KEM Hospital Research Centre, Pune for practical assistance during the trial (Charudatta Joglekar, Sonali Wagle, Vidya Mudliar, Komal Adawani, Rucha Wagh). We thank Dr. Arun Nanivadekar and Dr Mohan Gupte for their guidance. We are grateful to the participants of both the trials.

## Authorship

CY, ST, PY conceptualized and planned the study. SK, VK, HL, RL, PY and CY were involved in the trial conduct. DB, DR, NM performed all the laboratory measurements. Statistical analysis was done by TL and AB. CY, TL, SP and DB wrote the manuscript.

